# The acute phase inflammatory response as a key determinant of reduced lipid-associated antioxidant defenses in Chinese patients with major depressive disorder

**DOI:** 10.1101/2025.10.08.25337557

**Authors:** Tangcong Chen, Mengqi Niu, Yueyang Luo, Jing Li, Abbas F. Almulla, Yingqian Zhang, Michael Maes

## Abstract

**Background:** Major depressive disorder (MDD) is characterized by interacting neuro-immune, metabolic, and oxidative stress pathways. Here we examine the interactions between the acute phase (AP) response and serum lipids in Chinese MDD patients.

**Methods:** This case-control study assessed serum lipids and the AP response in 125 MDD patients and 40 healthy controls (HC), while controlling for metabolic confounders, including metabolic syndrome.

**Results:** We found an impaired lipid profile in MDD, characterized by reduced levels of high-density lipoprotein cholesterol (HDL), apolipoprotein (Apo) A1, chloromethyl phenylacetate (CMPA)ase activity, a reverse cholesterol transport (RCT) index, and increased ApoB/ApoA1 index. MDD was characterized by an AP response as conceptualized by lower serum albumin and transferrin and increased monomeric C-reactive protein (mCRP). The AP response was significantly and inversely associated with total cholesterol, HDL, low-density lipoprotein cholesterol, ApoA1, ApoB, lecithin cholesterol acyltransferase, CMPAase activity, and RCT index. After adjusting for the AP response, MDD diagnosis maintained significant independent associations with lower HDL-C, ApoA1, RCT, and higher ApoB/ApoA1 ratio. Multivariate (logistic) regression analyses confirmed that these lipid-inflammatory alterations strongly predict MDD diagnosis and clinical symptom severity. We found that around 81.5% of the MDD patients showed metabolic-inflammatory aberrations with a specificity of 82.1% and an area under the receiver operating characteristic (ROC) curve of 0.873. Lower ApoA1 emerged as a particularly robust protective biomarker, showing significant inverse relationships with affective and chronic fatigue severity scores.

**Conclusions:** These findings illuminate the complex interplay between a smoldering inflammatory response and lipid metabolism in many patients with MDD.

## 1. Introduction

Major depressive disorder (MDD) is a prevalent and disabling mental health condition with a well-documented neuro-immune, metabolic, and oxidative stress (NIMETOX) pathophysiology (Maes et al., 2025a; Maes et al., 2025b). There is also a strong comorbidity between MDD and metabolic disorders, metabolic syndrome (MetS), and cardiovascular disease (CVD) (Maes et al., 2025a). Growing evidence highlights the role of lipid metabolism dysregulation in mediating the link between the NIMETOX pathways and MDD (Jirakran et al., 2025a; Maes et al., 2025a). In particular, alterations in the lipid profile of individuals with MDD—including reduced high-density lipoprotein cholesterol (HDL), apolipoprotein (Apo) A1, paraoxonase 1 (PON1), lecithin–cholesterol acyltransferase (LCAT) activity, and consequently impairments in the reverse cholesterol transport (RCT)—have been consistently reported across diverse populations (Jirakran et al., 2025a; Jirakran et al., 2025b; Maes et al., 1994; Maes et al., 2025b; Maes et al., 1997; Persons and Fiedorowicz, 2016).

Moreover, lower serum levels of HDL, LCAT activity, PON1 activity, especially that of chloromethyl phenylacetate (CMPA)ase, and ApoA1 further compromise the anti-inflammatory and antioxidant properties of HDL, contributing to a pro-atherogenic state (Almulla et al., 2023; Maes et al., 1994; Sarandol et al., 2006). The lipid profile established in MDD frequently shows increased atherogenic potential as quantified using indices such as the Castelli risk index (total cholesterol/HDL), the triglyceride / HDL (atherogenic index of plasma) and ApoB/ApoA1 ratio (Jirakran et al., 2025a; Jirakran et al., 2025b; Maes et al., 1994; Nunes et al., 2015).

Beyond lipid-specific changes, MDD is also characterized by activation of the immune-inflammatory response system (IRS) with an acute phase (AP) response (or inflammatory response) (Maes, 1993; Maes et al., 2025a; Maes et al., 2025c). The latter is characterized by lowered levels of negative AP proteins, such as albumin and transferrin, and increased levels of positive AP proteins, such as monomeric C-reactive protein (mCRP) (Maes, 1993; Maes et al., 2025c; Maes et al., 1992; Maes et al., 1991).

This inflammatory milieu might not only exacerbate lipid dysfunction but might also reinforce a vicious cycle among the NIMETOX pathways (Maes et al., 2025a; Maes et al., 2025b; Wang et al., 2025). Moreover, it is important to note that ApoA1 functions as a key negative acute-phase protein, whose synthesis is suppressed during systemic inflammation (Khovidhunkit et al., 2004). Similarly, HDL levels decrease as part of the AP response, contributing to an impaired RCT capacity(Cabana et al., 1996). These impairments arise because RCT—a process mediated primarily by HDL, ApoA1 and LCAT, which together facilitate the efflux of cholesterol from peripheral tissues to the liver—is compromised under inflammatory conditions due to reduced expression of ATP-binding cassette transporters and downregulation of hepatic scavenger receptor BI (SR-BI)(Almulla et al., 2023; Wang et al., 1998). Additionally, PON1, an enzyme associated with HDL that confers antioxidant and anti-inflammatory properties, is also reduced in the context of inflammation and oxidative stress, further diminishing the atheroprotective functions of HDL (Morris et al., 2021). In MDD, these mechanisms are particularly relevant, as evidenced by consistent reports of inverse associations between HDL levels and immune-inflammatory biomarkers, underscoring the interplay between lipid metabolism and immune activation in this population (de Melo et al., 2017; Maes et al., 1997). Thus, the AP response might impact the lipid changes in MDD, leading to decreased HDL functionality and aberrations in RCT (Almulla et al., 2025a). However, the extent to which lipid abnormalities persist after controlling inflammatory markers in MDD has remained unclear.

Hence, in this study, we comprehensively assess the lipid profile associated with the AP response in a well-characterized cohort of MDD patients from Chengdu, China. We aim to clarify whether lipid alterations in MDD might be secondary to the AP inflammatory response or represent independent metabolic aberrations.

## 2. Methods

### 2.1. Participants

This cross-sectional study included a total of 165 participants, comprising 125 patients diagnosed with major depressive disorder (MDD) and 40 healthy controls (HC). Patients were recruited from the Psychiatric Center of Sichuan Provincial People’s Hospital in Chengdu, China. All participants were aged between 18 and 65 years. The diagnosis of MDD was confirmed using the Diagnostic and Statistical Manual of Mental Disorders, Fifth Edition (DSM-5) criteria, and a score above 18 on the 21-item Hamilton Depression Rating Scale (HAMD-21) was required for inclusion. Healthy controls were recruited from hospital staff, their family members, and acquaintances, and were matched to patients based on age, sex, education, and body mass index (BMI). Exclusion criteria for both groups included: (a) other major psychiatric disorders (e.g., bipolar disorder, schizophrenia, substance use disorders, autism spectrum disorders); (b) neurological (e.g., Parkinson’s disease, multiple sclerosis) or medical conditions (e.g., autoimmune diseases, rheumatoid arthritis, cancer, chronic inflammatory disorders, psoriasis); (c) recent infections, surgery, or use of immunomodulatory agents; and (d) pregnancy or lactation. Additionally, controls were excluded if they had a personal or family history of mood disorders, psychosis, or suicide.

All participants gave written informed consent prior to their involvement. The study was conducted in full compliance with international and Chinese ethical and privacy regulations and was formally approved by the Ethics Review Committee of the University of Electronic Science and Technology of China (approval no. 30005). It followed the ethical principles set forth in the Belmont Report and the guidelines of the International Council for Harmonisation Good Clinical Practice (ICH-GCP).

### 2.2. Clinical and Behavioral Assessments

All participants underwent a semi-structured interview conducted by a qualified physician to collect demographic and clinical characteristics, including age, gender, education, income, disease progression, medical history, and psychiatric history. The Mini International Neuropsychiatric Interview (M.I.N.I.) was used to screen and diagnose major mental disorders and antisocial personality disorder according to DSM-IV and ICD-10 criteria(Sheehan et al., 1998). On the same day, the same rater administered several clinical scales: the 21-item Hamilton Depression Rating Scale (HAMD) for depression severity(Hamilton, 1960), the Hamilton Anxiety Rating Scale (HAMA) for anxiety severity(Hamilton, 1959), the state version of the State-Trait Anxiety Inventory (STAI) for self-reported anxiety(Ferreira and Murray, 1983), and the FibroFatigue (FF) Scale for fibromyalgia and chronic fatigue syndrome symptoms(Zachrisson et al., 2002). A composite clinical severity index was constructed by summing the z-scores of HAMD, HAMA, STAI, and FF. The severity of adverse childhood experiences (ACEs) was evaluated with the Childhood Trauma Questionnaire-Short Form (CTQ-SF)(Bernstein et al., 2003), using the total score across its five subscales (emotional abuse, physical abuse, sexual abuse, emotional neglect, and physical neglect)(Vasupanrajit et al., 2024). Anthropometric measurements included waist circumference, height, and weight. Waist circumference was measured midway between the iliac crest and the lowest rib. BMI was calculated as weight (kg) divided by height (m) squared. A composite anthropometric index was derived as the sum of z-scores for BMI and waist circumference (z BMI+WC). MetS was defined based on the 2009 Joint Scientific Statement criteria(Alberti et al., 2009), requiring at least three of the following: elevated WC, triglycerides, blood pressure, fasting glucose, or reduced HDL cholesterol. The number of MetS criteria fulfilled was used as a MetS rank in subsequent analyses.

### 2.3. Assays

Between the hours of 06:30 and 08:00 a.m., a fasting venous blood sample of 30 mL was collected into serum tubes utilizing disposable syringes. The samples were subjected to centrifugation at a speed of 3500 rpm. Subsequently, the serum was carefully aliquoted into Eppendorf tubes and stored at a temperature of −80 °C until further analysis could be conducted. The measurements of albumin, transferrin, triglycerides (TG), total cholesterol (TC), Low-density lipoprotein cholesterol (LDL-C), high-density lipoprotein cholesterol (HDL-C), Apolipoprotein (Apo) A1, ApoB, ApoE, free cholesterol (FC), mCRP, (Chloromethyl)phenyl acetate (CMPAse) activity, and lecithin-cholesterol acyltransferase (LCAT) are listed in **Electronic Supplementary File (ESF), Table 1**. CRP was adjusted for the effects of age, sex, and BMI, and those residualized mCRP were used in the current study(Almulla et al., 2025b). The AP index was calculated using the formula z mCRP – z albumin – z transferrin (labeled AP index).

**Table 1.** Clinical, socio-demographic, and biochemical data of patients with major depressive disorder (MDD) and healthy controls (HC) All results are shown as mean (SD).

| Variables | HC (n=40) | MDD (n=125) | F/$\chi^2$ | df | p |
| --- | --- | --- | --- | --- | --- |
| Age (years) | 37.1(13.8) | 35.7 (12.1) | 0.37 | 1/163 | 0.542 |
| Gender (m/f) | 13/27 | 38/87 | 0.06 | 1 | 0.802 |
| BMI (kg/m<sup>2</sup>) | 23.52 (4.07) | 22.30 (3.6) | 3.58 | 1/163 | 0.060 |
| Waist circumference (cm) | 79.38 (11.73) | 78.27 (11.32) | 0.28 | 1/163 | 0.595 |
| Ranking MetS | 1.590 (1.46) | 1.42 (1.26) | 0.5 | 1/161 | 0.479 |
| Education years (years) | 13.88 (4.34) | 13.54 (3.33) | 0.26 | 1/163 | 0.613 |
| Metabolic syndrome (MetS) (No/Yes) | 29/10 | 102/22 | 1.17 | 1 | 0.355 |
| History of smoking (No/Yes) | 37/3 | 101/24 | 3.03 | 1 | 0.091 |
| Total HAMD | 0.9 (2.0) | 28.5 (5.2) | 1067.02 | 1/163 | <0.001 |
| Total HAMA | 1.1 (2.9) | 26.6 (8.6) | 338 | 1/163 | <0.001 |
| Total FF | 1.4 (2.9) | 29.9 (9.9) | 319.21 | 1/163 | <0.001 |
| OSOD | -1.518 (0.237) | 0.528 (0.491) | 640.21 | 1/153 | <0.001 |
| Total ACEs | 36.2 (9.4) | 47.5 (12.79) | 26.51 | 1/163 | <0.001 |
| Albumin (g/L) | 43.64 (1.99) | 41.18 (3.05) | 22.44 | 1/161 | <0.001 |
| Transferrin (mg/dL) | 258.76 (37.44) | 228.25 (38.66) | 18.79 | 1/162 | <0.001 |
| mCRP (z score) | -0.329 (0.941) | 0.103 (0.987) | 5.83 | 1/162 | 0.017 |
| AP index (z score) | -0.773 (0.993) | 0.243 (0.873) | 37.58 | 1/161 | <0.001 |

BMI: body mass index, HAMD: Hamilton Depression Rating Scale, HAMA: Hamilton Anxiety Rating Scale, FF: Fibro-Fatigue Scale, OSOD: overall severity of depression, ACEs: Adverse Childhood Experiences, mCRP: monomeric C-Reactive Protein (adjusted for age, sex, and body mass index), AP: acute phase response.

### 2.4. Statistics

Data are presented as means and standard deviations or estimated marginal means (SE). Group comparisons between MDD patients and controls were performed using Analysis of Variance (ANOVA) for continuous variables and chi-square tests for categorical variables. General Linear Models (GLM) were applied to compare lipid and atherogenic biomarkers between groups, adjusting for age, sex, and BMI where appropriate. False Discovery Rate (FDR) correction was applied for multiple comparisons. Multiple linear regression analyses were employed to predict clinical rating scale scores based on biomarkers and ACEs. Both manual and automated stepwise procedures were utilized in the process. The automated modeling implemented entry and removal thresholds of p = 0.05 and p = 0.07, respectively. The findings encompassed standardized beta coefficients, degrees of freedom, p-values, R², and F-statistics. The presence of heteroskedasticity was assessed utilizing both the White test and the modified Breusch–Pagan test methodologies. The assessment of multicollinearity was conducted through the examination of tolerance and variance inflation factors. Binary logistic regression models were employed to compare MDD with the control group. MDD served as the dependent variable, with controls as the reference group. The findings encompassed the Nagelkerke pseudo-R², Wald statistics accompanied by p-values, odds ratios with 95% confidence intervals, and regression coefficients (B) along with their respective standard errors (SE). The performance of the model was encapsulated through several metrics, including the area under the ROC (receiver operating characteristic) curve (AUC), the Gini index, the maximum Kolmogorov–Smirnov (Max K–S) values, as well as an assessment of overall fit. The classification accuracy, subjected to cross-validation, was evaluated utilizing linear discriminant analysis in conjunction with a 10-fold cross-validation methodology. Data transformations, including log10, square-root, rank-order, or Winsorizing, were implemented, as necessary.

## 3. Results

### 3.1 Features of MDD

**Table 1** presents the socio-demographic, clinical, and biochemical characteristics of the 165 study participants, including 125 patients with MDD and 40 healthy controls. No significant differences were observed between the two groups in terms of age, gender distribution, education, BMI, waist circumference, ranking of MetS, prevalence of MetS, or smoking history (all *p* >0.05). However, significant differences were found in several clinical and biochemical measures. MDD patients exhibited markedly higher scores on the total HAMD, HAMA, FF, and total ACE scores than controls. Biochemically, serum albumin and transferrin levels were significantly lower in the MDD group than in the control group. The residual mCRP data and the acute phase (AP) index were significantly increased in MDD patients, reflecting increased mCRP and lowered levels of both negative AP proteins.

### 3.2 Lipid profile of MDD

**Table 2** presents the results of GLM analysis comparing metabolic and lipid-related biomarkers between patients with MDD and HC, after adjusting for potential confounders (age, sex, BMI, MetS and smoking). Levels of HDL, ApoA1 and CMPAse were significantly lower in the MDD group compared to controls. Similarly, the reverse cholesterol transport marker RCT was also significantly reduced in MDD patients. The z composite scores ratios of z TG-HDL and z ApoB-ApoA1 were significantly higher in the MDD group, indicating a more atherogenic lipid profile. No significant differences were found between groups in triglycerides, total cholesterol, direct LDL, ApoB, ApoE, free cholesterol, and LCAT activity. After FDR correction, the following variables remained statistically significant: HDL (*p* < 0.001), ApoA1 (*p* < 0.001), the RCT index (*p* < 0.001), and the ApoB/ApoA1 ratio (*p* = 0.007).

**Table 2.** Differences in lipid profiles between major depressive disorder (MDD) and healthy controls (HC)

| Variables | HC (n=40) | MDD (n=125) | F | p | Partial Eta Squared |
| --- | --- | --- | --- | --- | --- |
| **TG (mmol/L)** | 1.393 (0.101) | 1.355 (0.056) | 0.205 | 0.651 | 0.001 |
| **TC (mmol/L)** | 4.849 (0.135) | 4.611 (0.074) | 2.372 | 0.126 | 0.015 |
| **direct LDL (mmol/L)** | 2.502 (0.095) | 2.433 (0.052) | 0.395 | 0.530 | 0.003 |
| **HDL (mmol/L)** | 1.588 (0.047) | 1.365 (0.026) | 16.07 | <0.001 | 0.093 |
| **ApoA1 (g/L)** | 1.665 (0.033) | 1.421 (0.018) | 40.947 | <0.001 | 0.207 |
| **ApoB (g/L)** | 0.939 (0.033) | 0.871 (0.018) | 3.061 | 0.082 | 0.019 |
| **ApoE (mg/dL)** | 3.812 (0.067) | 3.706 (0.037) | 1.86 | 0.175 | 0.012 |
| **Free cholesterol (mmol/L)** | 1.205 (0.042) | 1.125 (0.023) | 2.864 | 0.093 | 0.018 |
| **z LCAT (z score)** | -0.032 (0.162) | 0.010 (0.089) | 0.052 | 0.820 | 0.001 |
| **CMPAase (U/mL)** | 16.694 (0.540) | 15.444 (0.298) | 4.165 | 0.043 | 0.026 |
| **z RCT index (z score)** | 0.531 (0.143) | -0.167 (0.079) | 17.896 | <0.001 | 0.102 |
| **z TG – z HDL (z score)** | -0.257 (0.139) | 0.081 (0.077) | 4.412 | 0.037 | 0.027 |
| **z ApoB – z ApoA (z score)** | -0.392 (0.144) | 0.123 (0.079) | 9.621 | 0.002 | 0.058 |

All results are shown as estimated marginal means (SE), after adjusting for age, sex, BMI, smoking and the drug state (all df=1/157).
**TG:** triglycerides, **TC:** total cholesterol, **LDL:** low-density lipoprotein, **HDL:** high-density lipoprotein, **Apo:** apolipoprotein, **LCAT:** lecithin–cholesterol acyltransferase, **CMPA:** chloromethyl phenylacetate, **RCT:** reverse cholesterol transport.

**Electronic Supplementary File (ESF), Table 2** shows the measurements of the lipid variables in subjects with and without MetS (after covarying for age, sex, smoking and MDD). Total cholesterol, LDL, triglycerides, ApoB, ApoE, free cholesterol, Castelli risk index 1 and z ApoB – ApoA1 were significantly higher, and ApoA1, HDL and RCT index were significantly lower in MDD than in controls. We observed that 92 of the MDD patients were treated with antidepressants, 68 with benzodiazepines, 10 with mood stabilizers, and 44 with antipsychotic medications. We found that there were no significant effects of treatment with antidepressants, antipsychotics, and benzodiazepines on any of the lipids, even without FDR p correction. There was a significant effect of mood stabilizers on the LCAT activity index (F = 8.81, df = 1/153, p = 0.003). However, after p correction, these effects were no longer significant.

### 3.3. Effects of the AP response on the lipid profiles in MDD

As shown in **Table 3**, the AP response exerted a widespread and significant impact on multiple lipid parameters. Specifically, the AP index significantly suppressed triglycerides, total cholesterol, LDL, HDLc, ApoA1, ApoB, LCAT activity, CMPAase, and RCT. **Figure 1** shows the partial regression of serum ApoA1 on the AP response after adjusting for age, sex, and BMI. After adjusting for the AP response, the diagnosis of MDD showed significant effects on HDL, ApoA1, RCT index, and the ApoB/ApoA1 ratio. This indicates that the associations between MDD and CMPAase and z TG – z HDL disappeared after controlling for the AP index. These results suggest that many of the lipid abnormalities observed in MDD—particularly those related to RCT and ApoA1 levels—are strongly influenced by the accompanying AP response. Still, that part of the changes in those lipids in MDD is independent of the AP response.

**Table 3.** Effects of the acute phase (AP) index on the lipid profiles in major depressive disorder (MDD)

| Dependent Variable | Effects of the AP index |  |  | Effects of MDD |  |
| --- | --- | --- | --- | --- | --- |
|  | F | P | Sign | F | P |
| **TG** | 5.393 | 0.022 | . | 0.314 | 0.576 |
| **TC** | 12.518 | <0.001 | . | 0.002 | 0.968 |
| **LDL** | 6.518 | 0.012 | . | 0.244 | 0.622 |
| **HDLc** | 6.723 | 0.010 | . | 6.822 | 0.010 |
| **ApoA1** | 16.694 | <0.001 | . | 19.097 | <0.001 |
| **ApoB** | 9.402 | 0.003 | . | 0.115 | 0.735 |
| **ApoE** | 2.447 | 0.120 | NS | 0.342 | 0.559 |
| **Free cholesterol** | 0.041 | 0.840 | NS | 2.089 | 0.150 |
| **LCAT activity** | 5.118 | 0.025 | . | 1.349 | 0.247 |
| **CMPAase activity** | 6.415 | 0.012 | . | 0.669 | 0.415 |
| **RCT index** | 18.814 | <0.001 | . | 4.952 | 0.027 |
| **z TG – z HDL** | 0.001 | 0.976 | NS | 3.657 | 0.058 |
| **z ApoB – z ApoA1** | 0.118 | 0.732 | NS | 7.084 | 0.009 |

All results of GLM analysis with MDD as fixed factor and the AP index, age, sex, body mass index, smoking and drug state of the patient as covariates. Sign -: indicates a significant suppressant effect of the AP response on the lipid variables, NS: non-significant.
TG: triglycerides, TC: total cholesterol, LDL: low-density lipoprotein, HDL: high-density lipoprotein, Apo: apolipoprotein, LCAT: lecithin–cholesterol acyltransferase, CMPAase: chloromethyl phenylacetate-ase, RCT: reverse cholesterol transport.

**Figure 1.**
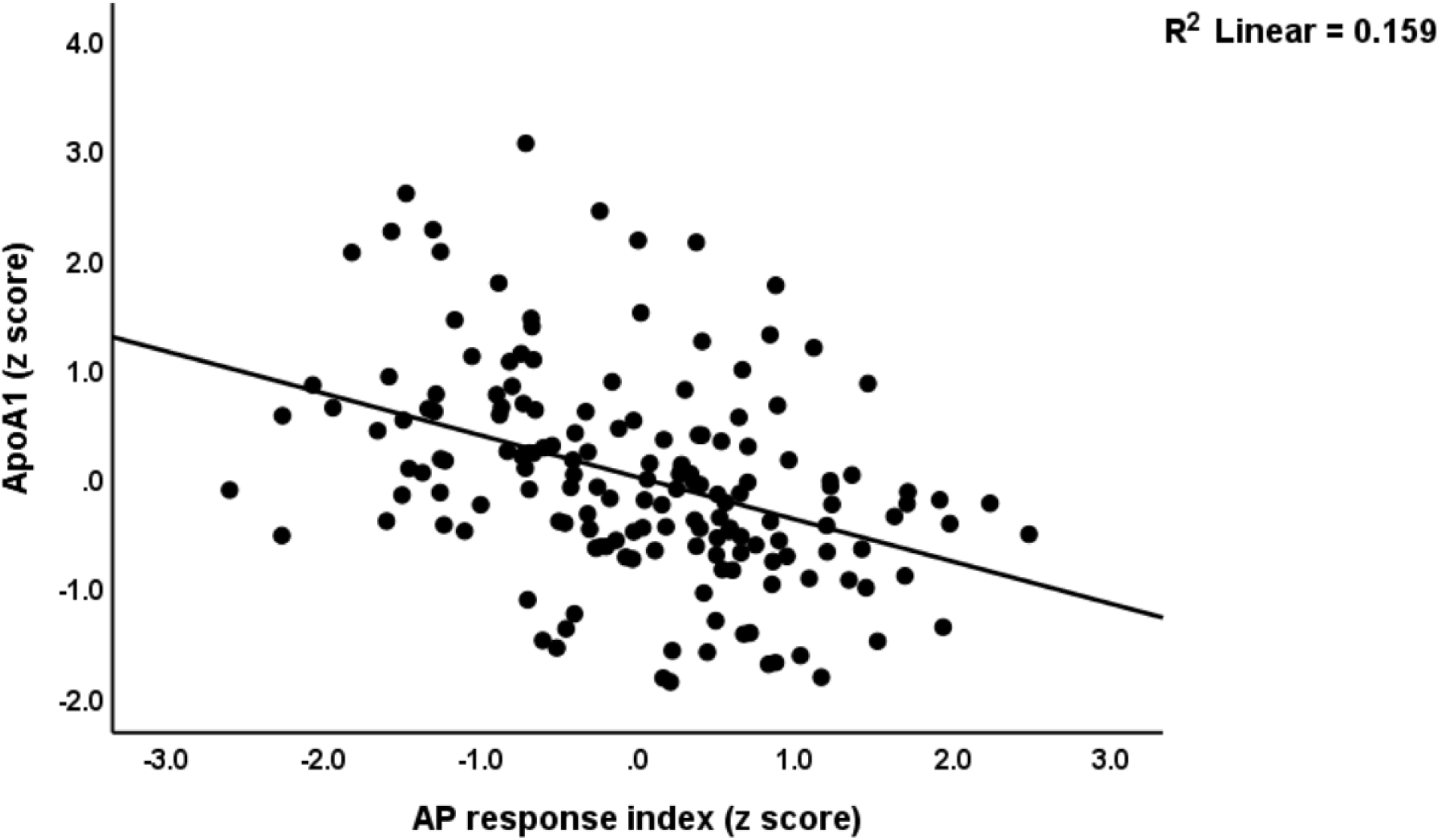
Partial regression of serum apolipoprotein A1 (ApoA1) on the acute phase (AP) inflammatory response after adjusting for age, sex, and BMI (p<0.001)

### 3.4 Multivariate discrimination of MDD from controls

**Table 4** displays the outcomes of binary logistic regression analyses performed to identify significant predictors that distinguish MDD patients from controls. Two models were constructed: one incorporating all subjects (with and without MetS), and another restricted to subjects without MetS. In the model utilizing all subjects, four variables emerged as significant independent predictors of MDD status (χ ^2^ = 117.56, df = 4, p < 0001, Nagelkerke = 0.515). The AP index and total cholesterol were found to elevate the likelihood of MDD, whereas ApoA1 and BMI served as protective factors. The overall accuracy of the model was 81.7% (sensitivity = 81.5% and specificity = 82.1%) and the area under the ROC curve was 0.873, SE = 0.023; Gini index = 0.746, max K-S-index = 0.654. **Figure 2** shows the ROC curve discriminating MDD from controls. Using LDA we found that the cross-validated diagnostic performance was sensitivity = 80.6% and specificity = 82.1%.

**Figure 2.**
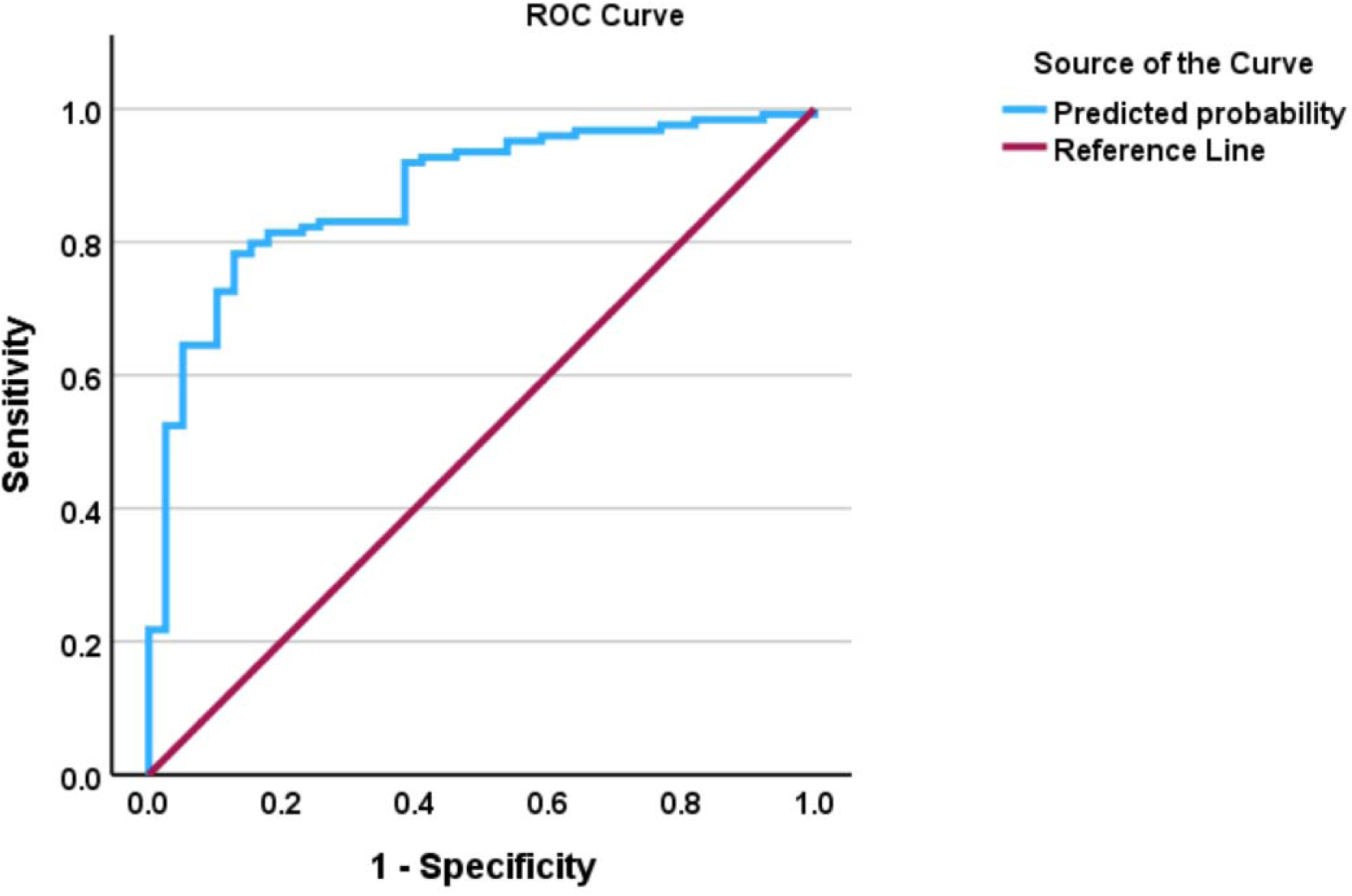
The receiving operating curve discriminating major depressive disorder from controls using apolipoprotein A, the acute phase inflammatory response, total cholesterol, and body mass index as explanatory variables.

**Table 4.**
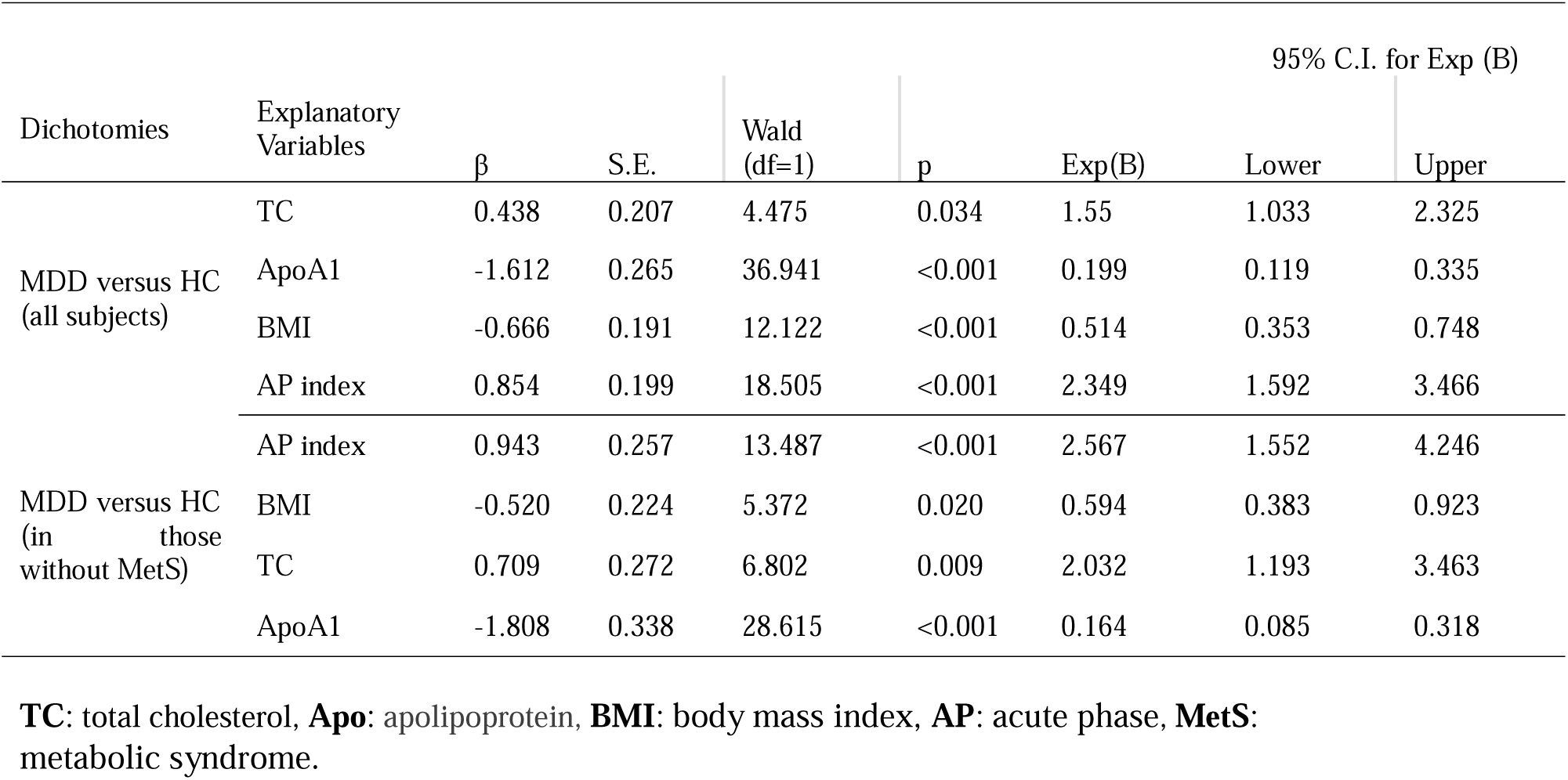
Results of binary logistic regression analysis examining the discrimination of major depressive disorder (MDD) and healthy controls (HC)

Analyzing the subset of subjects without MetS showed that the same predictors remained significant (χ ^2^ = 101.44, df = 4, p < 0001, Nagelkerke = 0.555). The overall accuracy of the model was 78.3% (sensitivity = 83.3% and specificity = 72.4%), and the area under the ROC curve was 0.890, SE = 0.023; Gini index = 0.780, max K-S-index = 0.617. Using Youden’s index (0.606) to delineate the optimal cut-off point, showed that 88.8% of the patients were correctly classified with a specificity of 73.4%. Using LDA we found that the cross-validated diagnostic performance was sensitivity = 78.4% and specificity = 79.3%.

### 3.5 Results of multiple regression analysis

**Table 5** presents a series of multiple regression models that quantify the key predictors of clinical symptom severity. All models were highly statistically significant and explained a substantial proportion of the variance (R^2^ ranging from 0.232 to 0.377). The severity of depressive symptoms, as measured by the total HAMD score (Model #1), was significantly predicted by the model (R^2^ = 0.321) with 4 key predictors: AP index, ACEs (both positively), ApoA1 and transferrin (both inversely). We found that 30.6% of the variance in the total HAMA score was significantly associated with ApoA1, albumin (both inversely), ACEs, and total cholesterol (both positively). The FF score (Model #3, R^2^ = 0.326 was predicted by ACEs (positively), ApoA1, albumin (both inversely), and male gender. We found that 32.2% of the variance in the STAl-state anxiety score was significantly associated with ACEs (positively), ApoA1 and BMI (inversely). Model #5 shows that 37.7% of the variance in the OSOD score was significantly predicted by ACEs, res mCRP (both positively), ApoA1 and transferrin (both inversely). **Figure 3** visually complements these findings by illustrating the partial regression of OSOD on ApoA1.

**Table 5.**
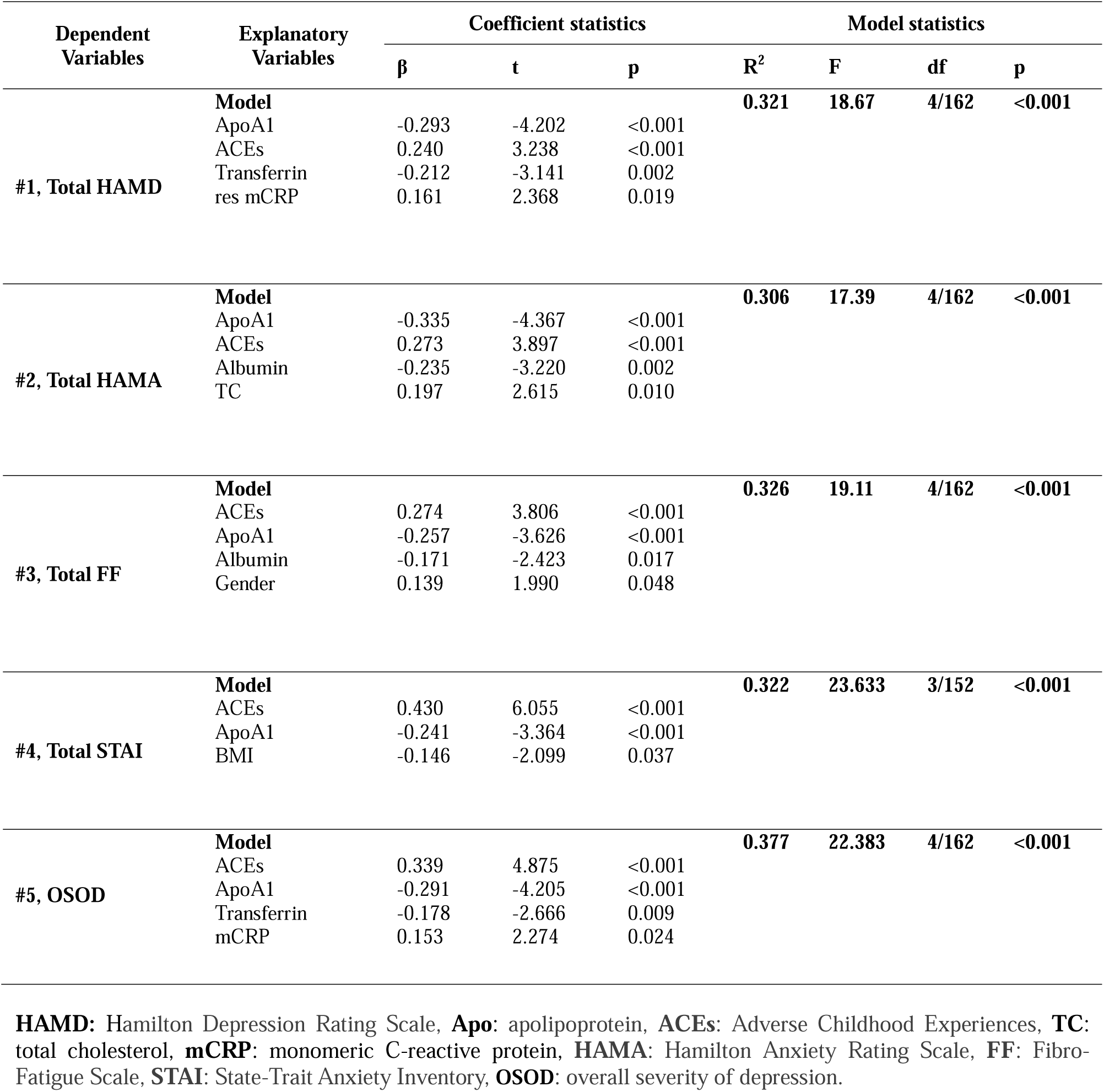
Results of Multiple regression analysis with clinical rating scale scores as dependent variables.

| Dependent Variables | Explanatory Variables | Coefficient statistics |  |  | Model statistics |  |  |  |
| --- | --- | --- | --- | --- | --- | --- | --- | --- |
| | | $\beta$ | t | p | R<sup>2</sup> | F | df | p |
| #1, Total HAMD | **Model** |  |  |  | **0.321** | **18.67** | **4/162** | **<0.001** |
|  | ApoA1 | -0.293 | -4.202 | <0.001 |  |  |  |  |
|  | ACEs | 0.240 | 3.238 | <0.001 |  |  |  |  |
|  | Transferrin | -0.212 | -3.141 | 0.002 |  |  |  |  |
|  | res mCRP | 0.161 | 2.368 | 0.019 |  |  |  |  |
| #2, Total HAMA | **Model** |  |  |  | **0.306** | **17.39** | **4/162** | **<0.001** |
|  | ApoA1 | -0.335 | -4.367 | <0.001 |  |  |  |  |
|  | ACEs | 0.273 | 3.897 | <0.001 |  |  |  |  |
|  | Albumin | -0.235 | -3.220 | 0.002 |  |  |  |  |
|  | TC | 0.197 | 2.615 | 0.010 |  |  |  |  |
| #3, Total FF | **Model** |  |  |  | **0.326** | **19.11** | **4/162** | **<0.001** |
|  | ACEs | 0.274 | 3.806 | <0.001 |  |  |  |  |
|  | ApoA1 | -0.257 | -3.626 | <0.001 |  |  |  |  |
|  | Albumin | -0.171 | -2.423 | 0.017 |  |  |  |  |
|  | Gender | 0.139 | 1.990 | 0.048 |  |  |  |  |
| #4, Total STAI | **Model** |  |  |  | **0.322** | **23.633** | **3/152** | **<0.001** |
|  | ACEs | 0.430 | 6.055 | <0.001 |  |  |  |  |
|  | ApoA1 | -0.241 | -3.364 | <0.001 |  |  |  |  |
|  | BMI | -0.146 | -2.099 | 0.037 |  |  |  |  |
| #5, OSOD | **Model** |  |  |  | **0.377** | **22.383** | **4/162** | **<0.001** |
|  | ACEs | 0.339 | 4.875 | <0.001 |  |  |  |  |
|  | ApoA1 | -0.291 | -4.205 | <0.001 |  |  |  |  |
|  | Transferrin | -0.178 | -2.666 | 0.009 |  |  |  |  |
|  | mCRP | 0.153 | 2.274 | 0.024 |  |  |  |  |

**HAMD:** Hamilton Depression Rating Scale, **Apo:** apolipoprotein, **ACEs:** Adverse Childhood Experiences, **TC:** total cholesterol, **mCRP:** monomeric C-reactive protein, **HAMA:** Hamilton Anxiety Rating Scale, **FF:** Fibro-Fatigue Scale, **STAI:** State-Trait Anxiety Inventory, **OSOD:** overall severity of depression.

**Figure 3.**
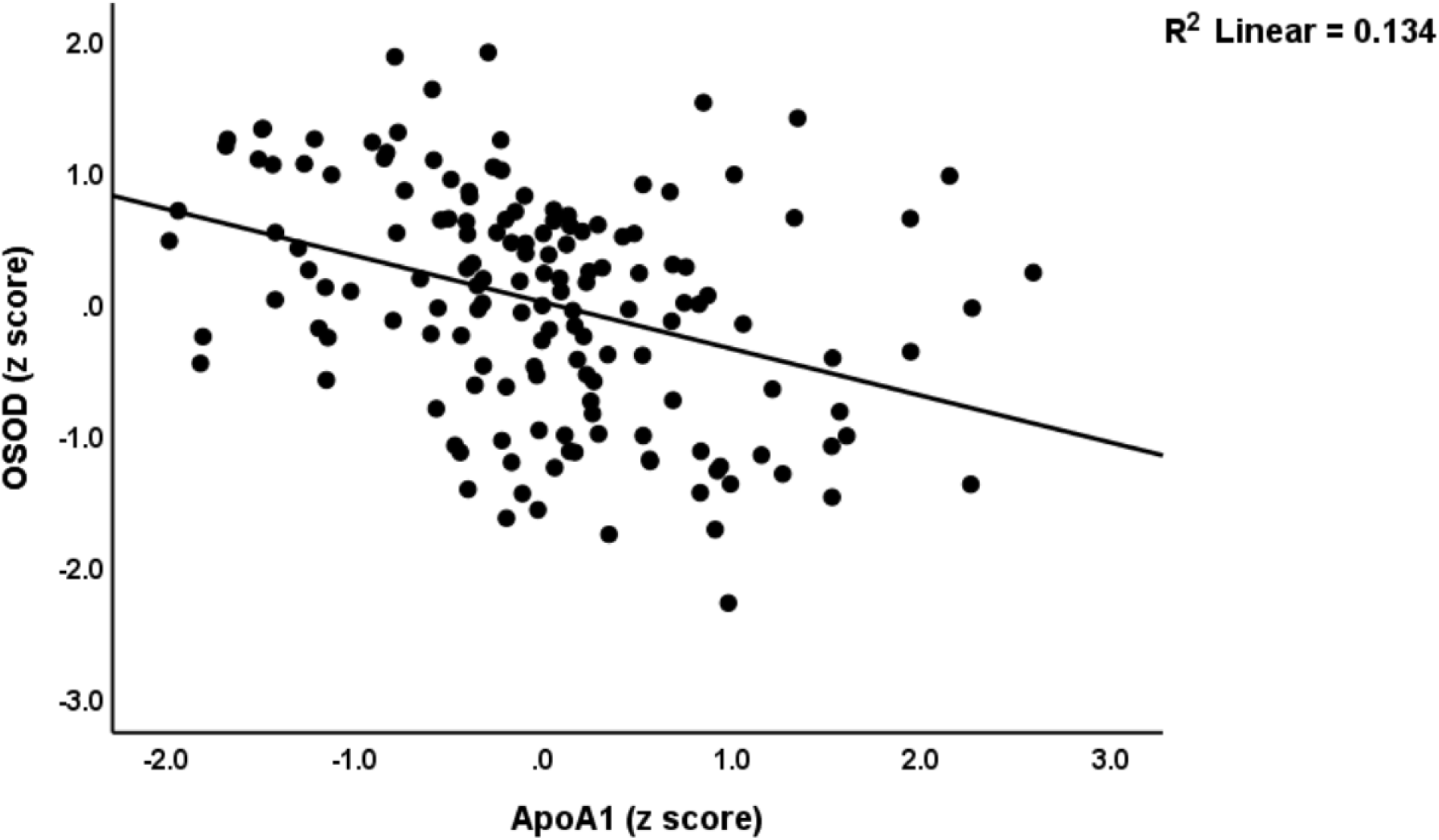
Partial regression of the overall severity of depression (OSOD) on serum apolipoprotein A1 levels after adjusting for age, sex, and education years (p<0.001)

## 4. Discussion

The present study provides a comprehensive analysis of lipid metabolism and the acute phase response in a well-characterized cohort of Chinese patients with MDD.

### 4.1 Serum lipids in MDD

The first major finding of this study is that a distinct pattern of lipid abnormalities characterized MDD, namely lower levels of HDL, ApoA1, and RCT index, alongside more modest elevations of z TG -z HDL, in patients compared to healthy controls. These results are largely consistent with those of a recent meta-analysis (Almulla et al., 2023; Jirakran et al., 2025a), which showed that MDD is associated with reduced HDL, ApoA1, and RCT capacity, as well as increased atherogenic indices. The persistence of similar lipid abnormalities in the current Chinese sample after excluding MetS patients highlights that the aberrations in lipid-associated biomarkers are intrinsic to the pathophysiology of MDD. Furthermore, the effect sizes of decreases in ApoA1 (19.9%) and the RCT index (11.5%) were much more important than the effects of MetS on these biomarkers (7.0% and 6.9%, respectively).

It was noted that the majority of lipid studies examined in the meta-analysis by Jirakran et al. did not account for MetS, BMI, overweight, or obesity (Jirakran et al., 2025a). Additionally, this lack of adjustment for metabolic variables caused considerable heterogeneity and bias. The significance is underscored by the high prevalence of MetS observed in the nations that analyzed lipid profiles included in this meta-analysis. For example, the incidence of MetS in the United States was 36.9% during 2015-2016 (Hirode and Wong, 2020), while the prevalence of obesity in US adults reached 40.3% from 2021 to 2023 (Emmerich et al., 2024)). In Belgium, where Maes et al. initially discovered increased atherogenicity in MDD (Maes et al., 1994), the prevalence of MetS was approximately 31.0% in 2018 (Leclercq et al., 2021). On the other hand, a population-based cohort in Chengdu (where we recruited our participants) showed a prevalence of 16.7% of MetS and 11.0% of obesity (using BMI > 28 kg/m2 in subjects of 49.1 (11.5) years old (Zhou et al., 2024), Our representative Chinese cohort showed a relatively low prevalence of MetS in both the MDD (22/124, 17.7%) and control (10/39, 25.6%) groups.

In order to more accurately scrutinize the effects of MetS on MDD-associated lipid measurement in countries with a high prevalence of MetS (for example, the USA population), a Thai study by Jirakran et al. (Jirakran et al., 2025b) recruited a selected sample of MDD patients, 50% of whom exhibited MetS and the other 50% did not. This study demonstrated that the results of lipids in MDD can only be assessed in study samples that control MetS. The inclusion of a significant number of patients with MetS results in an increase in heterogeneity and bias, which in turn obscures the actual lipid changes in MDD itself. This is due to the significant modification in lipids (e.g., HDL, total cholesterol, triglycerides) in MetS independently from MDD, which blurs the actual changes associated with MDD. Consequently, the results of most studies conducted in countries with a high prevalence of MetS or increased BMI (e.g., the United States and Europe) are challenging to interpret when metabolic variables are not controlled for.

All in all, the observation that significant lipid abnormalities were evident in the study sample with a numerically lower MetS prevalence (the current study) or after excluding those with MetS (Jirakran et al., 2025b) shows that the altered lipid profile is a feature of MDD pathophysiology itself, rather than being primarily attributable to concurrent MetS. In Chinese patients, the lipid profile of MDD differs from that of MetS. Both conditions are characterized by reduced ApoA1 levels (more significantly in MDD than in MetS), as well as diminished HDL and RCT indices; however, the atherogenicity indices are more prominent in MetS compared to MDD. Conversely, CMPAase activity is diminished in MDD, while no alterations in CMPAase are observed in MetS.

### 4.2 Association between the AP response and lipid alterations in MDD

The second major finding of this study is that our multivariate analyses demonstrated that the AP response, quantified by a composite index of mCRP, albumin, and transferrin, exerted a profound and widespread influence on the lipid profile. It should be noted that the AP index we used here reflects the positive (increases in mCRP) and the negative acute phase response (lowered albumin and transferrin) (Maes et al., 2025c). The AP index was most strongly associated with reductions in ApoA1, HDL, and RCT capacity (top three effect sizes), indicating that inflammation plays a central role in lipid dysregulation in MDD. These findings align with previous studies reporting inverse associations between inflammatory markers (e.g., IL-6, TNF-α) and HDL-related parameters in MDD (de Melo et al., 2017; Maes et al., 1997).

The observed suppression of RCT biomarkers is mechanistically explainable: during systemic inflammation, pro-inflammatory cytokines inhibit ApoA1 synthesis (Khovidhunkit et al., 2004), alter HDL composition (Cabana et al., 1996; Pruzanski et al., 2000), and downregulate key transporters such as ABCG1 and SR-BI, impairing cellular cholesterol efflux (Wang et al., 1998; Yvan-Charvet et al., 2010). Additionally, PON1 activity, which confers antioxidant capacity to HDL, is reduced under inflammatory conditions, further promoting a pro-atherogenic state (Morris et al., 2021; Rozenberg et al., 2003)). This lipid-inflammatory interplay is established in atherosclerosis and unstable angina (Mousa et al., 2022) and has also been observed in other conditions characterized by chronic inflammation, such as Long COVID (Bae et al., 2025). Such processes may convert HDL from an anti-inflammatory into an inflammatory compound (Morris et al., 2021).

Notably, after adjusting for the AP response, the MDD diagnosis retained significant independent effects on HDL, ApoA1, the RCT index, and the ApoB/ApoA1 ratio. This suggests that while inflammation drives a substantial portion of lipid dysfunction in MDD, certain metabolic disturbances may be intrinsic to the disorder. Genetic and nutritional factors, as well as other immune-specific or oxidative stress pathways, may offer additional mechanistic explanations (Maes et al., 1997; Morris et al., 2021).

### 4.3 Towards “region-specific strata in the immune–metabolic axis in MDD”

As reviewed above, comparative studies have revealed significant regional variations in lipid profiles between Western countries such as the USA and Asian countries such as China and Thailand. In addition, our Chinese cohort, which was recruited in Chengdu, exhibits a marked negative AP response pattern, as evidenced by significant decreases in negative AP reactants including albumin, transferrin, ApoA1, and HDL-C. In reality, the positive AP protein pCRP was not even elevated in this Chinese cohort (Maes et al., 2025c). A positive AP response dominated by elevated pCRP and haptoglobin levels in MDD is supported by a large-scale meta-analysis (Osimo et al., 2019). pCRP values may experience a significant increase (>10 mg/L in the case of autoimmune disorders and > 100 mg/L in infections) during acute inflammation, whereas MDD is characterized by no or only minor elevations (3-10 mg/dL) (Maes et al., 2025c). In fact, pCRP elevations in the lower serum range (3-10 mg/L) in MDD are more a biomarker of metabolic aberrations rather than inflammation (Almulla et al., 2025a; Almulla et al., 2025b). In addition, the inclusion of numerous individuals with MetS may be responsible for these minor increases in pCRP in MDD, a phenomenon that is particularly relevant to countries with a high prevalence of MetS/obesity. The latter effects are supported by the fact that BMI and MetS are the primary predictors of pCRP in MDD (Almulla et al., 2025a; Almulla et al., 2025b; Maes et al., 2025c).

In contrast, the modest more protracted inflammatory response in MDD is indicated by lower serum albumin, transferrin and ApoA1 levels and a modest increase in mCRP (this study, (Maes et al., 2025c)). The latter is indicative of a greater degree of chronic micro-inflammation, as observed in atherosclerosis and certain autoimmune disorders (Melnikov et al., 2023; Sproston and Ashworth, 2018; Thiele et al., 2014). Therefore, it seems that in Chengdu, China, there are significant changes in negative AP reactants and mCRP, indicating a mild smoldering inflammatory response associated with malnutrition and catabolism. Conversely, it appears that MDD in countries that exhibit an increased prevalence of MetS may additionally exhibit a more pronounced atherogenicity and pCRP response.

Additionally, Maes et al. (Maes et al., 2025b) discovered that the severity of MDD is significantly predicted by interactions between MetS/atherogenic indices and neurotoxicity cytokine profiles. This suggests that the relationship between immune biomarkers and MDD per se is challenging to interpret when a significant number of individuals with MetS are included in the study group. Nevertheless, in nations with a significant prevalence of MetS among their MDD study populations, the severity of MDD is undoubtedly influenced by the interplay between atherogenicity/MetS and immunological pathways (Jirakran et al., 2025b). However, these alterations do not represent the lipid fingerprint of MDD itself, but rather the interaction between comorbid metabolic variables and the pathophysiology of MDD.

To capture this geographic heterogeneity in lipid-immune responses in MDD, we propose the concept of a “region-specific immune–metabolic strata.” This framework helps explain why population studies in countries with a high incidence of MetS may show a positive AP-dominant profile that is driven by metabolic factors, whereas MDD in our Chengdu population exhibits a profile dominated by deficits in negative AP reactants and a very modest increase in mCRP. Importantly, such differences may perhaps be associated with distinct cardiovascular outcomes— e.g., coronary artery disease in countries with increased atherogenicity versus stroke in China (Collaborators, 2016; Farzadfar et al., 2011).

## Limitations

This study has several limitations that should be acknowledged. Firstly, the cross-sectional design prevents us from establishing firm causal relationships between the AP response and lipid abnormalities. Secondly, although we controlled for key confounders, residual influences from unmeasured variables such as diet, physical activity, and genetic factors cannot be ruled out. Additionally, despite excluding major comorbid conditions, the potential impact of undiagnosed or subclinical disorders remains. Future studies incorporating detailed oxidative stress profiling are needed to confirm our conclusions on NIMETOX pathway interactions in MDD. The outcomes of our multivariable regression analysis indicate that BMI was negatively correlated with the diagnosis of MDD and the severity of anxiety. The aforementioned “protective” effects are analogous to the previously noted “obesity paradox,” suggesting that elevated BMI mitigates the adverse consequences of infection and stroke by offering enhanced energy reserves (Lu et al., 2024; Yeo et al., 2023). These findings warrant replication in other investigations.

## Conclusions

In summary, our study demonstrates that MDD is associated with a particular lipid fingerprint, largely driven by an underlying AP response. However, deficits in HDL and ApoA1 persist even after accounting for the AP response, suggesting both inflammatory and non-inflammatory pathways contribute to this lipid profile in MDD. ApoA1 and HDL emerged as particularly robust lipid biomarkers in our analyses, inversely associated with depression and anxiety severity and serving as a protective factor in our diagnostic models.

Future research should develop targeted interventions to improve lipid profiles, and the immune system by metabolic interactions in MDD. Interventions aimed at increasing HDL and ApoA1 and attenuating the chronic smoldering AP response should be developed. Possible lipid-targeting interventions are statins (e.g., simvastatin), which have shown some pleiotropic anti-inflammatory effects (Salagre et al., 2016), PPAR-γ agonists (e.g., pioglitazone), which can modulate inflammation and lipid metabolism (Colle et al., 2017), or microbiota-modulating therapies (e.g., probiotics) that can influence both inflammation and lipid profiles (Goh et al., 2019). Future research should examine the causes of low HDL and ApoA1 levels which are independent of the AP response.

## Supporting information

Electronic Supplementary File

## Data availability

Data will be provided by the senior author (MM) upon a reasonable request.

## Ethical statement

This study was approved by the Ethics Committee of the Sichuan Provincial People’s Hospital, Affiliated Hospital of University of Electronic Science and Technology of China (approval no. 2024-203) and strictly followed ethical and privacy regulations.

## Author’s contributions

All the contributing authors have participated in the manuscript. Michael Maes, Mengqi Niu and Yingqian Zhang designed the study. Assays were performed by Yingqian Zhang, Tangcong Chen, Yueyang Luo, and Abbas F. Almulla. The first draft of this manuscript was written by Tangcong Chen, revised by Michael Maes and Yingqian Zhang, and edited by all other authors. Patients were recruited by Mengqi Niu and Jing Li. All authors have read and agreed to the published version of the manuscript.

---

**ESF, Table 1.**
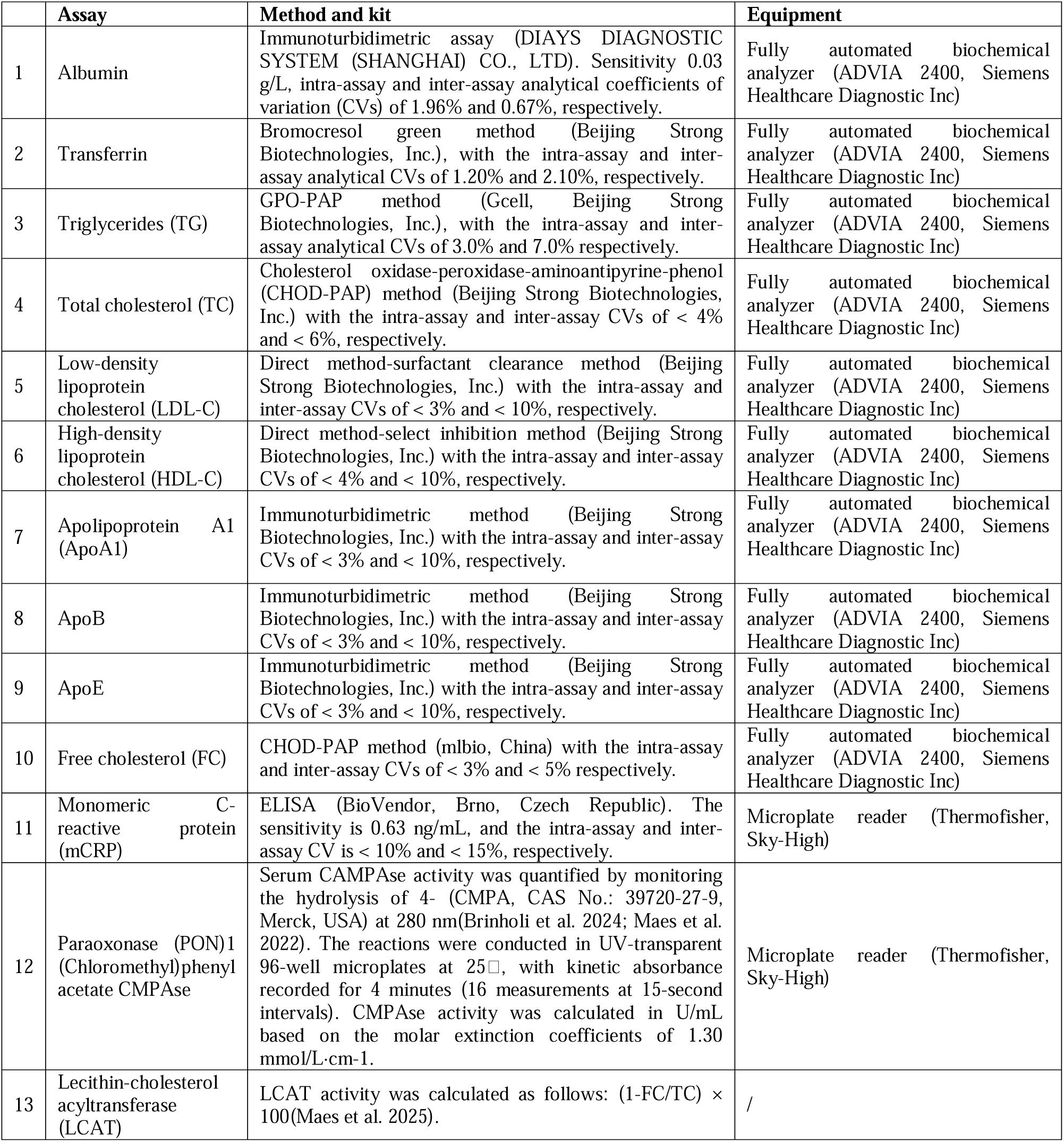
Methods used to assay the biomarkers in the present study.

**ESF, Table 2.**
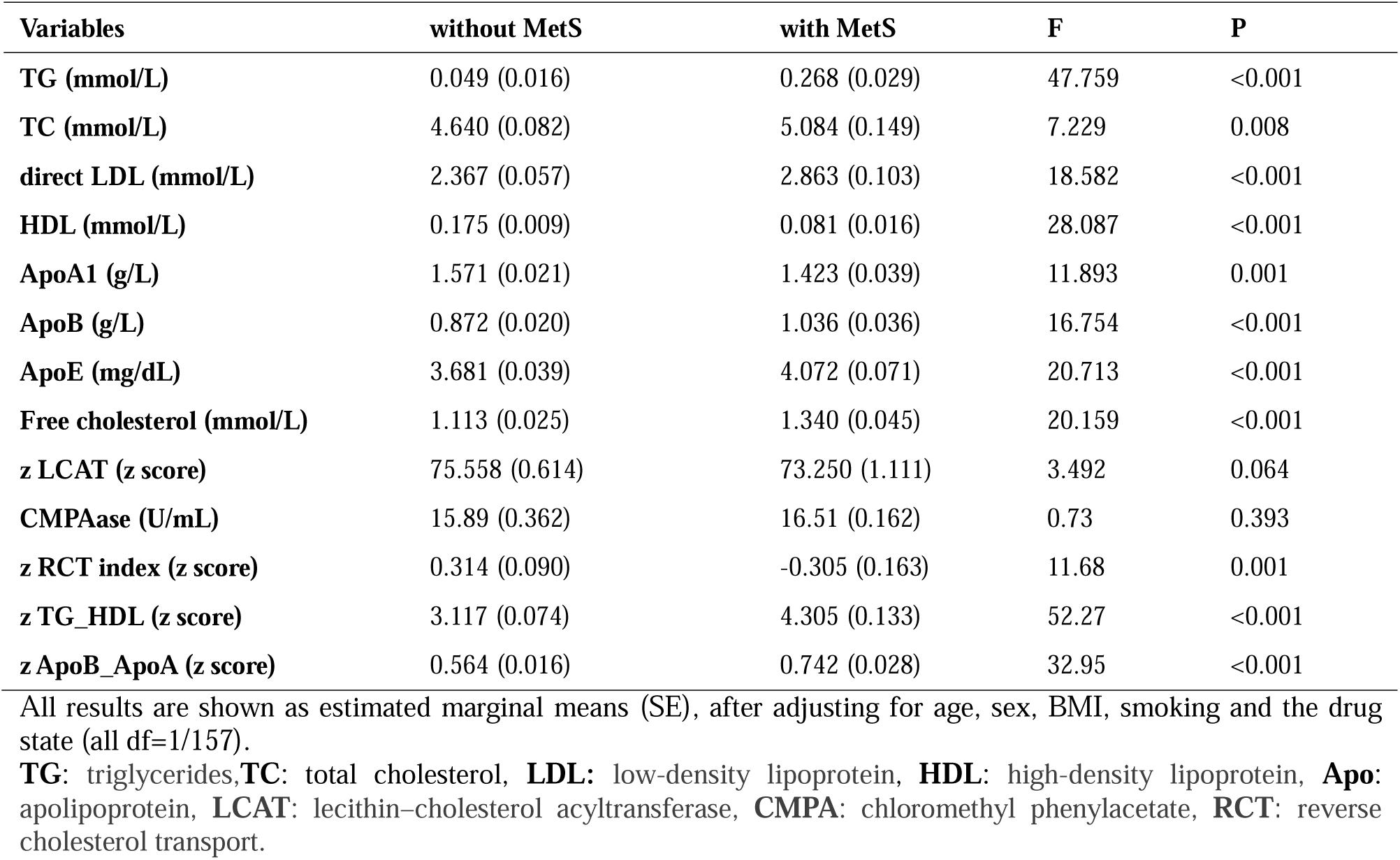
Differences in lipid profiles between people with and without metabolic syndrome (MetS)

