## Supplementary material for "The acute phase inflammatory response as a key determinant of reduced lipid-associated antioxidant defenses in Chinese patients with major depressive disorder": Electronic Supplementary File

**ELECTRONIC SUPPLEMENTARY FILE (ESF)**

**ESF, Table 1. Methods used to assay the biomarkers in the present study**

|  | **Assay** | **Method and kit** | **Equipment** |
| --- | --- | --- | --- |
| 1 | Albumin | Immunoturbidimetric assay (DIAYS DIAGNOSTIC SYSTEM (SHANGHAI) CO., LTD). Sensitivity 0.03 g/L, intra-assay and inter-assay analytical coefficients of variation (CVs) of 1.96% and 0.67%, respectively. | Fully automated biochemical analyzer (ADVIA 2400, Siemens Healthcare Diagnostic Inc) |
| 2 | Transferrin | Bromocresol green method (Beijing Strong Biotechnologies, Inc.), with the intra-assay and inter-assay analytical CVs of 1.20% and 2.10%, respectively. | Fully automated biochemical analyzer (ADVIA 2400, Siemens Healthcare Diagnostic Inc) |
| 3 | Triglycerides (TG) | GPO-PAP method (Gcell, Beijing Strong Biotechnologies, Inc.), with the intra-assay and inter-assay analytical CVs of 3.0% and 7.0% respectively. | Fully automated biochemical analyzer (ADVIA 2400, Siemens Healthcare Diagnostic Inc) |
| 4 | Total cholesterol (TC) | Cholesterol oxidase-peroxidase-aminoantipyrine-phenol (CHOD-PAP) method (Beijing Strong Biotechnologies, Inc.) with the intra-assay and inter-assay CVs of < 4% and < 6%, respectively. | Fully automated biochemical analyzer (ADVIA 2400, Siemens Healthcare Diagnostic Inc) |
| 5 | Low-density lipoprotein cholesterol (LDL-C) | Direct method-surfactant clearance method (Beijing Strong Biotechnologies, Inc.) with the intra-assay and inter-assay CVs of < 3% and < 10%, respectively. | Fully automated biochemical analyzer (ADVIA 2400, Siemens Healthcare Diagnostic Inc) |
| 6 | High-density lipoprotein cholesterol (HDL-C) | Direct method-select inhibition method (Beijing Strong Biotechnologies, Inc.) with the intra-assay and inter-assay CVs of < 4% and < 10%, respectively. | Fully automated biochemical analyzer (ADVIA 2400, Siemens Healthcare Diagnostic Inc) |
| 7 | Apolipoprotein A1 (ApoA1) | Immunoturbidimetric method (Beijing Strong Biotechnologies, Inc.) with the intra-assay and inter-assay CVs of < 3% and < 10%, respectively. | Fully automated biochemical analyzer (ADVIA 2400, Siemens Healthcare Diagnostic Inc) |
| 8 | ApoB | Immunoturbidimetric method (Beijing Strong Biotechnologies, Inc.) with the intra-assay and inter-assay CVs of < 3% and < 10%, respectively. | Fully automated biochemical analyzer (ADVIA 2400, Siemens Healthcare Diagnostic Inc) |
| 9 | ApoE | Immunoturbidimetric method (Beijing Strong Biotechnologies, Inc.) with the intra-assay and inter-assay CVs of < 3% and < 10%, respectively. | Fully automated biochemical analyzer (ADVIA 2400, Siemens Healthcare Diagnostic Inc) |
| 10 | Free cholesterol (FC) | CHOD-PAP method (mlbio, China) with the intra-assay and inter-assay CVs of < 3% and < 5% respectively. | Fully automated biochemical analyzer (ADVIA 2400, Siemens Healthcare Diagnostic Inc) |
| 11 | Monomeric C-reactive protein (mCRP) | ELISA (BioVendor, Brno, Czech Republic). The sensitivity is 0.63 ng/mL, and the intra-assay and inter-assay CV is < 10% and < 15%, respectively. | Microplate reader (Thermofisher, Sky-High) |
| 12 | Paraoxonase (PON)1 (Chloromethyl)phenyl acetate CMPAse | Serum CAMPAse activity was quantified by monitoring the hydrolysis of 4- (CMPA, CAS No.: 39720-27-9, Merck, USA) at 280 nm(Brinholi et al. 2024; Maes et al. 2022). The reactions were conducted in UV-transparent 96-well microplates at 25℃, with kinetic absorbance recorded for 4 minutes (16 measurements at 15-second intervals). CMPAse activity was calculated in U/mL based on the molar extinction coefficients of 1.30 mmol/L·cm-1. | Microplate reader (Thermofisher, Sky-High) |
| 13 | Lecithin-cholesterol acyltransferase (LCAT) | LCAT activity was calculated as follows: (1-FC/TC) × 100(Maes et al. 2025). | / |

**ESF, Table 2.** Differences in lipid profiles between people with and without metabolic syndrome (MetS)

| **Variables** | **without MetS** | **with MetS** | **F** | **P** |
| --- | --- | --- | --- | --- |
| **TG (mmol/L)** | 0.049 (0.016) | 0.268 (0.029) | 47.759 | <0.001 |
| **TC (mmol/L)** | 4.640 (0.082) | 5.084 (0.149) | 7.229 | 0.008 |
| **direct LDL (mmol/L)** | 2.367 (0.057) | 2.863 (0.103) | 18.582 | <0.001 |
| **HDL (mmol/L)** | 0.175 (0.009) | 0.081 (0.016) | 28.087 | <0.001 |
| **ApoA1 (g/L)** | 1.571 (0.021) | 1.423 (0.039) | 11.893 | 0.001 |
| **ApoB (g/L)** | 0.872 (0.020) | 1.036 (0.036) | 16.754 | <0.001 |
| **ApoE (mg/dL)** | 3.681 (0.039) | 4.072 (0.071) | 20.713 | <0.001 |
| **Free cholesterol (mmol/L)** | 1.113 (0.025) | 1.340 (0.045) | 20.159 | <0.001 |
| **z LCAT (z score)** | 75.558 (0.614) | 73.250 (1.111) | 3.492 | 0.064 |
| **CMPAase (U/mL)** | 15.89 (0.362) | 16.51 (0.162) | 0.73 | 0.393 |
| **z RCT index (z score)** | 0.314 (0.090) | -0.305 (0.163) | 11.68 | 0.001 |
| **z TG_HDL (z score)** | 3.117 (0.074) | 4.305 (0.133) | 52.27 | <0.001 |
| **z ApoB_ApoA (z score)** | 0.564 (0.016) | 0.742 (0.028) | 32.95 | <0.001 |

All results are shown as estimated marginal means (SE), after adjusting for age, sex, BMI, smoking and the drug state (all df=1/157).

**TG**: triglycerides,**TC**: total cholesterol, **LDL:** low-density lipoprotein, **HDL**: high-density lipoprotein, **Apo**: apolipoprotein, **LCAT**: lecithin–cholesterol acyltransferase, **CMPA**: chloromethyl phenylacetate, **RCT**: reverse cholesterol transport.
